# A cross-sectional study investigating the prevalence and causes of vision impairment in Northwest Portugal using capture-recapture

**DOI:** 10.1101/2021.07.06.21260069

**Authors:** Pedro Lima Ramos, Rui Santana, Ana Patrícia Marques, Inês Sousa, Amândio Rocha-Sousa, Antonio Filipe Macedo

## Abstract

**Objectives:** The aim of this study was to estimate the prevalence and causes of vision impairment (VI) in Portugal.

**Setting:** Information about people with VI was obtained from Primary Care Centres, blind association (ACAPO) and from hospitals (the PCVIP-study) in the Northwest of Portugal during a period spanning years 2014-2015. Causes of VI were obtained from hospitals.

**Participants:** Administrative and medical records of people with visual acuity in the better seeing eye of 0.5 decimal (0.30logMAR) or worse and/or visual field less than 20 degrees were investigated. Capture-recapture with log-linear models was applied to estimate the number of individuals missing from lists of cases obtained from available sources.

**Primary and secondary outcome measures:** Log-linear models were used to estimate the crude prevalence and the category specific prevalence of VI.

**Results:** Crude prevalence of VI was 1.97% (95%CI=1.56-2.54), and standardized prevalence was 1% (95%CI=0.78-1.27). The age-specific prevalence was 3.27% (95% CI=2.36-4.90), older than 64 years, 0.64% (95%CI=0.49-0.88), aged 25-64 years, and 0.07% (95%CI=0.045-0.13), aged less than 25 years. The female-to-male ratio was1.3, that is, higher prevalence amongst females. The five leading causes of VI were Diabetic Retinopathy, Cataract, Age-related Macular Degeneration, Glaucoma and Disorders of the Globe.

**Conclusions:** The prevalence of VI in Portugal was within the expected range and in line with other European countries. A significant number of cases of VI might be due to preventable cases and, therefore, a reduction of the prevalence of VI in Portugal seems possible. Women and old people were more likely to have VI and, therefore, these groups require extra attention. Future studies are necessary to characterize temporal changes in prevalence of VI in Portugal.

**Strengths and limitations of this study:**

- Medical records and registers of people with vision impairment were used to determine the number of cases in these sources.
- Data from 3 sources (lists) with records about people with vision impairment were combined using log-linear models to determine the number of “uncaptured” cases.
- Capture-recapture methods were used to determine the prevalence of vision impairment in the Northwest Portugal.
- Capture-recapture methods to compute prevalence are more accurate than pure case counting from lists and more affordable than cross-sectional studies.
- A limitation of the current study was the low completeness, that is, the number of cases captured compared with the number of uncaptured cases.

## Introduction

Vision impairment (VI) leads to a significant loss of quality of life mostly due to activity limitations, loss of independence and difficulties to find jobs [1-6]. Because VI leads to a significant burden it is important to have regular vigilance (estimates) of cases of VI so that the quality of eye care and events such as diseases that may be leading to more cases of VI can be detected, evaluated, monitored and, eventually, vision loss can be prevented [7, 8]. One example of initiatives based on prevalence was VISION 2020 - an action of World Health Organization and the International Agency for the Prevention of Blindness, whose aim was to prevent and monitor VI and promote vision rehabilitation worldwide [9]. Recent estimates indicate that VI remains a significant health problem in Europe; although, in some countries reliable and updated information is lacking [9].

In 2020, in Western Europe, it has been estimated that there were 15 million people with moderate or severe VI [9]. However, the prevalence of VI and the methodology for the estimation varies significantly from country to country. For example, a population-based study conducted in Denmark in 2016 defined VI as best corrected visual acuity worse than 20/40 (0.3 logMAR) in the better-seeing eye. The study involved people aged 20 to 94 years and found a prevalence of 0.4% (95% CI, 0.2–0.7) [10]. A very different estimation with significantly different values was performed France in 2005 [11], in this study VI was self-reported and the prevalence was 1.95%. In 2007 and for the population aged 50 years or older, a study from Hungary reported a prevalence of 0.5% (95% CI, 0.2–0.7) for severe VI and 5.1% (95% CI, 4.3–5.9) for moderate VI [12]. These numbers are often hard to compare due to different age categories included and recruitment methods used; although, they point to differences among European nations.

Differences in prevalence of VI within Europe, as summarized in **Table 1**, may be due to not only study design but also, for example, due to differences in disease prevalence. VI and blindness in Western Europe is mostly linked to age-related eye diseases, in Germany, for example, that corresponds to 70% of all cases of blindness [13]. In Scotland, the leading causes of VI are Age-related Macular Degeneration (AMD), Glaucoma, Diabetic Retinopathy (DR), Myopic Degeneration, and Optic Atrophy [14]. Differences in disease prevalence and disease severity are associated with factors such as prevention and access to treatments [8]. Inequalities in accessing treatments can be seen even within a single country such as Portugal where unequal access to anti-VGEF injections has been detected [15]. This makes it important to investigate prevalence and causes of VI as detailed as possible at national and regional levels. One method to study prevalence when cross-sectional studies of the population are unavailable is called capture-recapture (CR).

**Table 1:** Prevalence of visual impairment in European countries. SVI= severe visual impairment; MVI=moderate visual impaiment; VA= visual acuity.

| Country | Year | Sample | VI definition | Prevalence |
| --- | --- | --- | --- | --- |
| **Denmark[10]** | 2016 | 3826 participants aged 20-94 years old from the Danish General Suburban Population Study | The best corrected visual acuity worse than 20/40 in the better-seeing eye | 0.4% (95% CI, 0.2–0.7) |
| **Hungary[12]** | 2017 | 105 clusters of 35 people 50 years of age or older | SVI-VA < 6/60–3/60<br>MVI- VA < 6/18–6/60 | SVI, 0.5% (95% CI, 0.2–0.7)<br>MVI, 5.1% (95% CI, 4.3–5.9) |
| **France[11]** | 2005 | Random stratified sample of 359 010 French citizens | Self-reported visual impairment | 1.95% |
| **Spain[16]** | 2014 | 213 626 participants aged ≥15 years from a 2008 Spanish Survey | Distance and near visual impairment distinguished by applying a questionnaire | Near visual impairment - 1.89%<br>Distance visual impairment - 1.89% |
| **Germany[7]** | 2019 | Population-based cohort study in Germany concerning 14687 adults aged 35 to 74 | Acuity below 0.3 in the better-seeing eye | 0.37% (95% CI, 0.28–0.49) |
| **Iceland[1]** | 2008 | Random sample of 1045 citizens of Reikjavik aged 50 or more years | Bilateral VI - best corrected visual acuity VA < 6/18 or visual field of > or = 5 degrees and < 10 degrees around the fixation point in the better eye | 0.96% (95% CI, 0.37–1.55) |
| **UK[17]** | 2002 | 14600 participants aged 75 years and older | Binocular visual acuity <6/18 | 12.4% (95% CI, 10.8–13.9) |

CR methods have been used to estimate the prevalence of several eye conditions [18-26]. CR methods is a methodology that can overcome the problem of cases that are never captured by, for example, registers for the visual impaired [27-29]. For a detailed description on how to use CR methods we recommend reading our review about the method [30]. Due the lack of information about the prevalence of VI in Portugal, we conducted a study with CR methods using data from different sources. The aim of this study was to estimate the prevalence and the main causes of VI in Portugal using CR methods.

## Methods

Information about people with VI was obtained from different sources in the Northwest of Portugal during a period spanning years 2014-2015. The geographical coverage included 42 municipalities from two provinces: Minho, population density = 241.1 inhabitants/km^2^ and Douro Litoral, population density = 742.4 inhabitants/km^2^ as reported by national CENSUS 2011 [31]. The study was conducted in accordance with the tenets of the Declaration of Helsinki, approved by the local ethics committees of the participating hospitals and by the ethical committee for Life Sciences and Health of the University of Minho, Ref. SECVS-084/2013; data protection process numbers 9936/2013 and 9793/ 2017.

Possible sources of patients with visual acuity in the better eye of 0.5 decimal (0.30logMAR) or worse and/or visual field less than 20 degrees were investigated [32]. The first source were Primary Care Centres that were used for list L1. This list contained subjects that applied, for example, for medical certificates of VI. According to the Portuguese law people with a level of impairment of 60% or more are entitled to, amongst other, tax exemption, completely free health care or early retirement [33]. As an example, to get a degree of 60% or more from vision only, one eye must have no measurable acuity and the other can have acuity up 0.2 decimal [34] Although, for this certificate all types of impairments can be combined, e.g. vision and motor impairments, to reach the final score. Because of that, cases mapped as having VI were analysed and only those with field or acuity matching the inclusion criteria were included in this study. The second source used was an association for the visually impaired named ACAPO and their records were used in the second list L2. To be member of ACAPO people must be visually impaired (low vision or blind).

The third list L3 was obtained from the PCVIP-study, a hospital-based study whose aim was to determine prevalence, causes and costs of VI in Portugal [35-37].The study gathered demographic, clinical, and socioeconomic information of people with VI. Participants were selected among patients attending ophthalmologists’ appointments at four Portuguese public hospitals: Hospital de Braga, Hospital Senhora da Oliveira-Guimarães, Hospital de Santa Maria Maior-Barcelos and Centro Hospitalar e Universitário de São João -Porto. The initial database included people with acuity 0.5 decimal (0.30logMAR) or worse. To make compatible with the definition of VI in the ICD9 [32] and with the acuity in the other lists only cases with visual acuity below 0.3 decimal (0.5 logMAR) in the better-seeing eye were used for the estimation of prevalence.

All lists had variables that allowed assessment of repeated cases by using a string or combination of strings that form identifiers or “tags”. Information available included: initials, date of birth, sex and municipality. The list from the hospitals also included information about the cause of VI. An example of a tag could be JS130519802 where JS are the initials (first and last name), 13051980 is the date of birth (13-05-1980) and the last digit (2) defines sex – 2 is a female in the example given. By matching the identity strings (tags) of the three lists, it was possible to ascertain the number of individuals present in all three lists and the number of individuals present at any combination of two lists.

### Application of the CR method

To be used in CR lists need to be obtained at approximately the same time, or based on different sources that represent approximately the same population [38]. In addition, to obtain reliable results with CR certain assumptions need to be meet: 1) the sources of lists are independent - this implies that the probability of a subject being in both list A and list B equals the product between the probability of being in A alone and the probability of being in B alone [39], 2) the probability of association within each source (catchability) is equal for all individuals - the probability may vary from one list to another, or be constant overall [39, 40], 3) the population is closed (no births, deaths or migrants). These assumptions are restrictive and, when applied to medical conditions, are unlikely to be strictly followed. Log-linear models are one way of handling, for example, lists that are not completely independent [39].

Log-linear models were applied to estimate the number of individuals missing from all three lists [40]. Log-linear models result from the application of Poisson regression models to **Table 2** which summarizes all possible capture history for all cases listed. The capture histories are illustrated in **Figure 1**. The logarithm of the count in each cell of the table is modelled as a linear function with terms indicating the presence or absence in the lists and terms modelling possible pairwise dependences between lists. Log-linear models compute the expected value of *n*_*ijk*_, that is, the expected value for the number of individuals with capture history *(i j k)*. For example, according to **Table 2** or **Figure 1**, *n*_111_ = 13. When there are three lists and there is no simultaneous dependence amongst the three but a dependence between any possible pair of lists the equation for the log-linear model is:

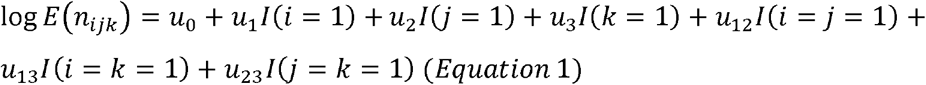

**Table 2:** Number of individuals presenting each possible capture history.

| L1 | L2 | L3 | Freq |
| --- | --- | --- | --- |
| 1 | 1 | 1 | 13 |
| 1 | 1 | 0 | 38 |
| 1 | 0 | 1 | 39 |
| 0 | 1 | 1 | 59 |
| 1 | 0 | 0 | 118 |
| 0 | 1 | 0 | 768 |
| 0 | 0 | 1 | 4161 |
| 0 | 0 | 0 | *x* |

**Figure 1.** Venn diagram representing the overlap between the three lists.

In Equation 1, *I*(*i*= 1) stands for the function that assigns 1 to the capture histories (1 *j k*) and 0 to all the others. The parameter *u*_12_ models the dependence between lists 1 and 2, and *u*_13_ the dependence among lists 1 and 3 and equivalent to other pairs. By using Equation 1 we can compute the expected value for the number of individuals with capture history (*i j k*), that is, *E*(*n*_*ijk*_).

The parameters of the model can be computed using the R package Rcapture.[41, 42]. For example, using Equation 1, the software can compute the expected value for the number of individuals with capture history (1 1 1), that is, *E*(*n*_111_) and compares the model estimation with the true value of *n*_111_, in our case the real intersection would be 13 cases (Figure 1). This process can be repeated for all capture histories except for (0 0 0) which are the missing cases (hidden population). Then the null hypothesis can be tested, that is, “the observed cell counts are equal to the estimated cell counts”. In other words, “does the model fits the data well”? – that is given by the Qui-square goodness of fit test. After modeling all possible parameters, the Rcapture software can use Equation 1 to compute *E*(*n*_000_) and that is the size of the hidden population. The final estimate for *E*(*n*_000_) is given by the best model that should pass the Qui-square goodness of fit test and should be the one with the lowest value of AIC.[43] A review of the method with an intuitive link to a video (https://www.youtube.com/watch?v=aiSKgIc_8vk) is given in a review by Bird and King (2018)[44] and in our previous publication[30].

After choosing the best possible model for our data we obtained an estimate of the size of the hidden population and consequently we estimate of the number of individuals with VI. The same procedure was used to compute category specific prevalence according to age and sex. Within each category several models were applied to sub-lists obtained from the main lists.

### Patient and Public Involvement

Members of the Blind Association ACAPO were involved in the design and data collection of this study.

## Results

The total number of inhabitants in the geographical area covered by the current study was 3 010 964. The list from Primary Care Centres (L1) had 208 cases (52% females) with a mean age of 60 years (SD=18.93). The list with the cases from ACAPO (L2) had 878 cases (43% females) with a mean age of 54 years (SD=18.0). The list from the Hospitals (L3) had 4272 cases (58% females) with a mean age of 74 years (SD=18.0). The Venn diagram in **Figure 1** shows the intersection between lists obtained from comparisons between identity strings. **Figure 1** shows that, for example, 39 individuals were in L3 and L1 and were not in L2; 13 individuals were in all three lists; 4161 individuals were only in L3. Table 2 provides the possible capture histories and the number of individuals with that history. For example, a subject has a capture history (1 1 0) when she or he was in L1 and L2 but not in L3.

Log-linear models admitting possible list dependence scenarios were applied to model the counts in **Table 2**. The model is expected to estimate the value of *x* (**see also Table 2**) that corresponds the number of individuals with capture history (0 0 0). That is, the size of the hidden population or the number of individuals not captured by any of the three lists. The estimate of total number of people with VI (N) was given by the expression: N= *x*+13+38+39+59+118+768+4162, the value of N changes from model to model because the estimates obtained to the unknown *x* value. All possible list dependence scenarios were considered resulting in seven models summarized in **Table 3**. Code used to implement these models in R Statistics (v3.6.1), package Rcapture [41, 42] is provided in **Appendix A**.

**Table 3:**
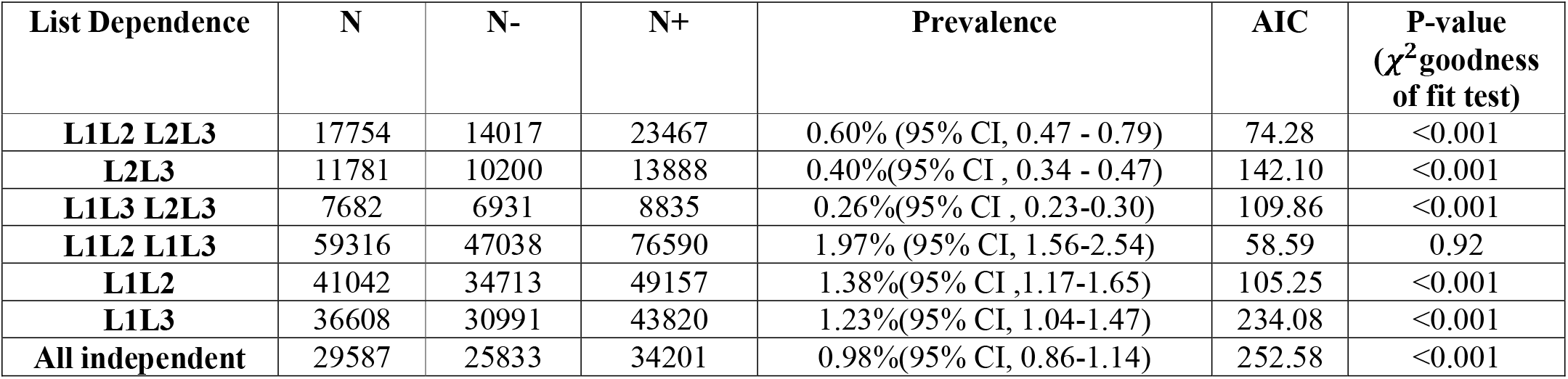
All possible log-linear models and resulting prevalence estimates. When we write, for example, L1L2 we are indicating that the model assumed dependence between L1 and L2. N- and N+ represent lower and upper estimates of N according to a 95% confidence interval. P-values test the hypothesis of the model fitting well (a value above 0.05 is indicative that the difference between the model predictions and the data is not statistically significant) the data and the Akaike’s Information Criteria (AIC) is a criterion to choose between models by considering a balance between the number of fitted parameters and the maximum likelihood.

The list dependence scenario L1L2 and L1L3 generated a model fitting the data well, Qui-Square goodness of fit test = 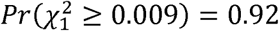 and corresponds to the model:

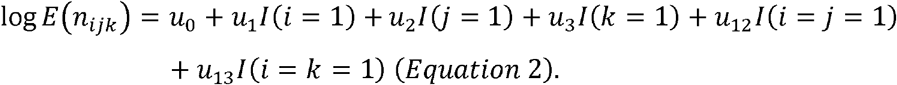

According to the model given by Equation 2 the crude prevalence of VI as estimated in this study was 1.97% (95% CI, 1.56-2.54). The standardize prevalence was 1% (95% CI, 0.78-1.27).

Completeness, that is, the proportion of the population with VI that has been captured by our three lists, assuming that the size N of the population with VI was 59 316, was 9%. Completeness was computed using the formula below, in the formula, *n*_100_ is the number of cases with capture history (1 0 0) and the meaning is the same for all other parcels such as *n*_010_ in the denominator of the fraction.

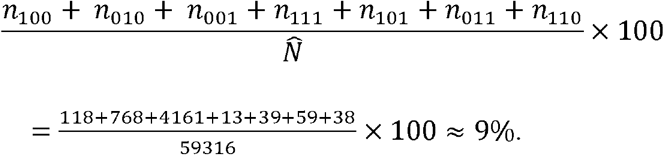

**Table 4** summarizes the category specific prevalence according to age and according to sex. To run new log-liner models for each category we divided the initial lists according to the desired categories. Subsamples for each category were used to generating new Venn diagrams. Log-linear models for each subsample were set as given in **Table 3**, that means seven different dependency scenarios for each, for example, age category. The best model was chosen using Qui-Square goodness of fit tests and AIC.

**Table 4:** Age-specific prevalence and sex-specific prevalence. P-values test the hypothesis of the model fitting well (a value above 0.05 is indicative that the difference between the model predictions and the data is not statistically significant) the data and the Akaike’s Information Criteria (AIC) is a criterion to choose between models by considering a balance between the number of fitted parameters and the maximum likelihood.

| Age-specific prevalence |  |  |  |
| --- | --- | --- | --- |
| Age | Prevalence | AIC | P-value ($\chi^2$ goodness of fit test) |
| <25 | 0.07% (95% CI, 0.045-0.13) | 22.37 | 0.94 |
| 25-64 | 0.64% (95% CI, 0.49-0.88) | 53.94 | 0.77 |
| >64 | 3.27% (95% CI, 2.36-4.90) | 50.43 | 0.82 |
| Sex-specific prevalence |  |  |  |
| Sex | Prevalence | AIC | P-value ($\chi^2$ goodness of fit test) |
| Male | 1.67% (95% CI, 1.32-2.19) | 28.85 | 1 |
| Female | 2.20% (95% CI, 1.65-3.08) | 28.58 | 1 |

**Table 5** summarizes the distribution of causes of VI. This information was available from L3 (from the hospitals), causes were classified according to the ICD9. DR was the most common cause of VI with 31% (95% CI, 29 - 32) of the cases in L3, followed by Cataract 15% (95% CI, 14 - 17), AMD 14% (95% CI, 13 - 15), Glaucoma 10% (95% CI, 9 - 11) and Disorders of the globe (DG) 5% (95% CI, 4 - 6).

**Table 5:** Summary of the causes of visual impairment, bold font highlights the mains causes for each age group.

| **Causes of visual impairment** | **Less than 25 years**<br>*N (%)* | **25-64 years**<br>*N (%)* | **More than 64 years**<br>*N (%)* | **All ages combined**<br>*N (%)* |
| --- | --- | --- | --- | --- |
| Age-related Macular Degeneration | -- | 29 (4%) | 550 (17%) | 579 (14%) |
| Cataract | 10 (7%) | 58 (7%) | 592 (18%) | 660 (15%) |
| Chorioretinal inflammations, scars, and other disorders of choroid | 1 (%) | 28 (3%) | 17 (1%) | 46 (1%) |
| Cornea | 1 (1%) | 54 (7%) | 44 (1%) | 99 (2%) |
| Disorders of the globe | 5 (3%) | 79 (10%) | 129 (4%) | 213 (5%) |
| Optic nerve disorders | 11 (8%) | 40 (5%) | 49 (1%) | 100 (2%) |
| Other retinal disorders | 4 (3%) | 18 (2%) | 36 (1%) | 58 (1%) |
| Diabetic Retinopathy | -- | 196 (25%) | 1110 (33%) | 1306 (31%) |
| Glaucoma | 3(2%) | 66 (8%) | 354 (11%) | 423 (10%) |
| Retinal detachments and defects | 6 (4%) | 37 (5%) | 72 2%) | 115 (3%) |
| Others | 102 (71%) | 189 (24%) | 382 (11%) | 673 (16%) |
| Total - sum of rows | 143 (3%) | 794 (19%) | 3335 (78%) | 4272 (100%) |

## Discussion

The current study investigated the prevalence and causes of VI in the Northwest of Portugal. Crude estimates of prevalence point that 2 out of 100 inhabitants of the Portuguese north-western population suffer from VI. Category-specific prevalence by age and by sex revealed higher prevalence among older people and among women. The top causes of VI included DR and Cataract, information about causes of VI was available only from cases detected at hospitals.

Prevalence of VI for the sample of the general Portuguese population was within the expected values. Our results are in line with those reported in neighbour countries such as Spain [16]. This was an expected result because both countries have similar demographics and health systems. Our results are also in line with a French study reporting a prevalence of VI of 1.95% [11]. A study from Iceland reported a prevalence of 0.96% (95% CI, 0.37–1.55) [1] that is similar to our study if we consider the standardized prevalence instead of the crude prevalence. Another study conducted in 2000 in Copenhagen, urban Denmark, also found a value for prevalence close to 1% [45]. In contrast, a study from 2016 in rural Denmark found a prevalence of 0.4% (95% CI, 0.2–0.7) [10] which is similar to what has been reported in Germany 0.37% (95% CI, 0.28–0.49) [7]. Recent studies show that the incidence of VI in countries like Germany has been reducing and, therefore, more recent studies are likely to report lower prevalence of VI than older studies [46]. One possible explanation for slightly higher values of prevalence of VI in our study in Portugal can be the prevalence and incidence of, for example, diabetes and DR [47, 48]. In other words, some European countries seem better at preventing vision loss from common eye diseases such has DR and removing it from top cause of VI, while in Portugal at the time of our study DR was still the top cause of VI [49]. In other parts of Europe such as Hungary prevalence of VI was higher than our study, here VI affected more than 5% of the population [12]. In short, prevalence of VI in Portugal was similar to neighbour countries, but slightly higher than in countries with, possibly, better preventive mechanisms of vision loss. Our results point that is possible to reduce the prevalence of VI in Portugal, the exact strategies can be inspired from European countries reporting lower prevalence of VI.

VI was more common among elderly people, it increased from about 7 out of 10,000 in the population under 25 years to 60 out of 10,000 in the age range 25-64 years and about 300 out of 10,000 in the population with 64 or more years, these findings are in line with other studies [7]. A study in Denmark found that VI was 9 times more prevalent amongst people with more than 64 years than amongst people in the age range 20-64 years [45]. Our results for the older population are also in line with the estimates from a recent meta-analysis estimating the prevalence of VI in people 55 years or older in European countries. The study that included data from Portugal, estimated an overall prevalence of VI for those above 55 years close to 2.75% [50, 51]. For age under 25 years, prevalence of VI in our study was low and in line with several other studies [52-54]. For example, our results were similar to data available from Sweden, in 1997 the age-specific prevalence of VI as 10.9/10,000 amongst people under the age of 19 years [54]. A more recent study from China that investigated VI amongst pre-school children also found a similar prevalence [55]. There was a good agreement between our results and similar studies, small differences might be due, among other factors, to temporal changes in prevalence of VI and the age-range criteria.

Prevalence of VI among females was 1.3 times higher than the prevalence among males, this result is in line with the trend reported in a recent meta-analysis covering European countries [50]. These results are also consistent with studies from Germany [7], VI among females was 1.4 times higher than among males, and from Spain [16], prevalence amongst females was 1.7 times higher than amongst males. The female-to-male ratio is expected to vary from 1.1 is Sub-Saharan Africa to 1.25 in Europe [56]. Causes for this female-to-male ratio above 1 are likely to include factors such as gender inequalities in access to health care [57].

The top two causes of VI in our study were DR and Cataract. Information about causes of VI in our study was available from hospitals and that might increase the frequency of cases with treatable diseases such as the top two causes. The main causes of VI in Europe are diverse [58] and we provide a summary of some studies in **Table 6** [7, 14, 45, 59, 60]. Studies compiled in **Table 6** show that, for example, DR was the top cause of VI only in our study. We speculate that the main reason was that when our study was conducted the preventive effects of DR screening were not yet visible in Portugal and, therefore, the number of cases of VI caused by DR was high [47]. This contrasts with other countries such as Germany or Denmark where DR appears down in the list of main causes of VI. Probably preventive DR measures were implemented earlier than in Portugal [47]. While in some studies DR remains in second place as cause of VI [61] it seem that the trend is to go further down in the list [62, 63]. Our second cause of VI, Cataract, has also been reported as important cause of VI in Denmark, Canada and the UK [10, 64, 65]. We believe that, for example, in Denmark, the high number of cases of Cataract causing VI was due to the inclusion criteria with acuity 20/40. In many countries this is also the criteria to undergo cataract surgery. In our study we believe that a considerable percentage of cases of VI caused by Cataract was due to long queues for surgery at the time of our study [66].

**Table 6:** Causes of visual impairment in 6 European countries, including Portugal.

| **This study** | **Denmark[45]** | **Scotland[14]** | **Italy[59]** | **Poland[60]** | **Germany[7]** |
| --- | --- | --- | --- | --- | --- |
| Diabetic retinopathy | Cataract | Age related macular degeneration | Cataract | Age related macular degeneration | Age related macular degeneration |
| Cataract | Age related macular degeneration | Glaucoma | Myopia | Cataract | Glaucoma |
| Age related macular degeneration | Diabetic retinopathy | Cataract | Age related macular degeneration | Amblyopia | Diabetic retinopathy |
| Glaucoma | Myopic degeneration | Diabetic retinopathy | Diabetic retinopathy | Diabetic retinopathy | Corneal Disease |
| Cornea | Other retinal causes | Myopia | **not available** | Cornea | Genetic illness |

In this study we used capture-recapture models to investigate prevalence of VI. Some models showed high quality of fit which gives credibility to the prevalence values that we obtained. The best models were the ones with the list dependences Primary Care Centres/Hospitals and Primary Care Centres/Blind Association. Internal validity of the models was assessed using Qui-Square goodness of fit tests and AIC. Only the models assuming the list dependence scenario Primary Care Centres/Hospitals and Primary Care Centres/Blind Association passed the Qui-Square goodness of fit test. The Primary Care Centres/Hospitals dependence is understandable because medical certificates of VI require a report from an ophthalmologist that, most likely, is the assistant physician at the hospital. The Primary Care Centers/Blind Association dependence is explained by the fact that the Blind Association recommends their members to get a medical certificate of VI. It was impossible to assess in the detail the external validly of our model that computed the hidden population. However, when we compare prevalence of VI for people above 55 years reported by a recent systematic analysis in European countries (prevalence 2.75%)[50] with the estimates in the current study for people with more than 64 years, 3.27% (95% CI, 2.36-4.90), it seems like our estimates are accurate. The fact that completeness was about 9% is a limitation of our study, to solve this we needed more information from Primary Care Centres. This limitation may be addressed in future studies with better standardized digital records that allow more efficient anonymous data sharing.

In conclusion, the results of the current study showed that prevalence of VI in Portugal was within the expected range and in line with other neighbour countries. A significant number of cases of VI detected was due to preventable causes, in other words, a reduction of cases of VI in Portugal is possible with improved access to eye care and effective diseases monitorization. In addition, basic and comprehensive vision rehabilitation is necessary to support people with VI [67]. Future studies are necessary to characterize temporal changes and the efficacy of public health measures such as DR screening at reducing prevalence of VI.

## Data Availability

Upon request from the corresponding author

## Contributorship

Contributed equally to this work Pedro Lima Ramos and Antonio Filipe Macedo.

Roles: Conceptualization, Data curation, Formal analysis, Investigation, Methodology, Software, Validation, Visualization, Writing – original draft, Writing – review & editing.

Rui Santana, roles - Conceptualization, Funding acquisition, Methodology, Project administration, Resources, Supervision, Validation, Writing – review & editing

Ana Patricia Marques roles - Data curation, Investigation, Validation, Visualization, Writing – review & editing. Inês Sousa, roles - Methodology, Supervision, Validation, Writing – review & editing. Amandio Rocha-Sousa - Conceptualization, Methodology, Supervision, Validation, Writing – review & editing.

## Funding

This study was supported by FCT (COMPETE/QREN) grant reference PTDC/DPT-EPI/0412/2012 in the context of the Prevalence and Costs of Visual Impairment in Portugal: a hospital-based study (PCVIP-study). PLR is funded by FCT (COMPETE/QREN) grant reference SFRH/BD/119420/2016. AFM if founded by the faculty of Health and Life Sciences at Linnaeus University.

## Competing interests

None of the authors have competing interests to disclosure.

## Acknowledgments

Authors report on behalf of the Portuguese visual impairment study group (PORVIS-group): António Filipe Macedo, PhD, Principal Investigator; Department of Medicine and Optometry Linnaeus University Kalmar, Sweden and Vision Rehabilitation Lab Centre/Department of Physics and Optometry University of Minho Braga, Portugal. Amandio Rocha-Sousa, MD, PhD, FEBO; Marta Silva, MD Ophthalmologist; Sara Perestrelo, MD Ophthalmologist; João Tavares-Ferreira, MD Ophthalmologist; Ana Marta Oliveira, research coordinator; Department of Surgery and Physiology, Faculty of Medicine University of Porto and/or Ophthalmology Department: Centro Hospitalar e Universitário de São João. Cristina Freitas, MD Ophthalmologist; Keissy Sousa, MD Ophthalmologist; Ricardo Leite, MD Ophthalmologist; José Carlos Mendes, MD Ophthalmologist; Andreia Braga Soares, MD Ophthalmologist; Rui Carneiro Freitas, MD Ophthalmologist; Department of Ophthalmology, Hospital de Braga. Pedro Reimão, MD Ophthalmologist; Marco Vieira, MD Ophthalmologist; Joel Monteiro, MD Cardiologist; Department of Ophthalmology, Centro Hospitalar de Alto Ave, Guimarães. Natacha Moreno, MD, Ophthalmologist; Department of Ophthalmology, Hospital Sta Maria Maior, Barcelos. Gary Rubin, PhD (project adviser); UCL-Institute of Ophthalmology, London, UK. Ana Patricia Marques, PhD; Rui Santana, PhD; Principal Investigator; National School of Public Health, NOVA University of Lisbon, Portugal. Laura Hernandez-Moreno, PhD candidate; Pedro Lima, PhD candidate; Low Vision and Visual Rehabilitation Lab, Department and Center of Physics – Optometry and Vision Science, University of Minho, Braga, Portugal.

## Data sharing statement

Raw data and MATLAB code to link the lists is available from the corresponding author upon request.

## Notes

### Competing Interest Statement

The authors have declared no competing interest.

### Author Declarations

The study was conducted in accordance with the tenets of the Declaration of Helsinki, approved by the local ethics committees of the participating hospitals and by the ethical committee for Life Sciences and Health of the University of Minho, Ref. SECVS-084/2013; data protection process numbers 9936/2013 and 9793/ 2017.

### Summary of Updates

We have now changed the title and the manuscript to restrict our findings to the target region where the study was performed. We have now improved the description of the model used in methods sub-section "Application of the CR method". We also added information about internal validity of the model below Table 3 and in discussion below Table 5.

